# Prognostic significance of circulating exosomal PD-L1 in various malignant tumors: A systematic review and meta-analysis

**DOI:** 10.1101/2024.01.20.24301560

**Authors:** Wentao Li, Qian Cui, Ting Ge, Shuangcui Wang, Dong Wang, Guixin He, Jianchun Yu

**Affiliations:** Department of Oncology, First Teaching Hospital of Tianjin University of Traditional Chinese Medicine, Tianjin, China; National Clinical Research Center for Chinese Medicine Acupuncture and Moxibustion, Tianjin, China; Graduate School of Tianjin University of traditional Chinese Medicine, Tianjin, China; Medical Experiment Center, First Teaching Hospital of Tianjin University of Traditional Chinese Medicine, Tianjin, China; National Clinical Research Center for Chinese Medicine Acupuncture and Moxibustion, Tianjin, China

**Keywords:** Circulating exosomal PD-L1, Prognostic significance, Immune checkpoint inhibitors, Meta-analysis

## Abstract

Although the prognostic significance of exosomal PD-L1 (exoPD-L1) has been previously reported, its value is still controversial. For the first time, we conducted a meta-analysis to systematically evaluate the prognostic value of exoPD-L1 in various types of cancer. The pooled hazard ratio (HR) and 95% confidence interval (CI) in these studies were used to explore the relationship between these indexes and overall survival (OS), recurrence-free survival (RFS), and progression-free survival (PFS). Utilizing the NewcastleLJOttawa Scale (NOS), the quality of the listed studies was assessed. Heterogeneity was explored by subgroup analyses. Begg’s and Egger’s tests assessed publication bias. This meta-analysis included 11 trials involving 964 cancer cases. The pooled results indicate that high-level pre-treatment exoPD-L1 in circulation was associated with worse OS (HR = 2.10, 95% CI 1.51–2.91, P < 0.001), RFS (HR = 1.67, 95% CI 1.18–2.37, P < 0.01) and PFS (HR = 3.49, 95% CI 2.60–4.68, P < 0.001) compared to those with low-level pre-treatment exoPD-L1. However, high fold changes in circulating exoPD-L1 after receiving immune checkpoint inhibitors (ICIs) were correlated with significantly superior OS (HR = 0.19, 95% CI 0.10–0.38, P < 0.001) and PFS (HR = 0.35, 95% CI 0.23–0.52, P < 0.001). Through this meta-analysis, we found that pre-treatment with high levels of exoPD-L1 is associated with a poor prognosis. However, a high fold change in circulating exoPD-L1 following immunotherapy was correlated with a superior prognosis. ExoPD-L1 may have important clinical significance for assessing the prognosis of cancer patients.

## Introduction

The success of cancer immunotherapy highlights the role of immune checkpoint molecules in promoting the progression and growth of malignant tumors.^1^ Among these molecules, the programmed cell death protein 1 (PD-1)/programmed death-ligand 1 (PD-L1) signaling pathway mediates immune tolerance under pathological conditions.^2^ When tumor antigens are recognized by T-cell, the release of IFN-γ triggers the expression of PD-L1 on tumor cells, ^3–4^ which then binds to PD-1 on the surface of activated T cells. This leads to the dephosphorylation of the T-cell receptor and its co-stimulatory receptor, CD28, inhibiting antigen-driven T cell activation.^5^ As a result, cancer cells evade anti-tumor immunity, leading to tumor survival, growth, and invasion.

Recently, immune checkpoint inhibitors (ICIs) have shown efficacy against various types of cancer, such as melanoma, non-small cell lung cancer, and kidney cancer. Although numerous clinical studies have confirmed the potential of ICIs in treating several types of cancer, only 20% of patients have responded to such therapies, ^6–7^ and some individuals have experienced severe disease progression or fatal toxicity. ^8^

It is essential to investigate readily available and repeatable biomarkers that can predict clinical outcomes in cancer patients treated with ICIs. Currently, PD-L1 expression and tumor mutational burden are recommended biomarkers. ^9^ However, several studies have reported that some patients with positive PD-L1 expression may not benefit from ICIs. ^10–11^

Exosomes, as the primary component of extracellular vesicles, are biologically active lipid bilayer vesicles naturally released via various normal and tumor cells. Emerging evidence suggests that cancer-derived exosomes are enriched in immunosuppressive molecules, including PD-L1.^12^ Recently, it has been found that malignant glioma cells can produce extracellular vesicles containing PD-L1, which participate in tumor progression. Exosomal PD-L1 (exoPD-L1) is involved in immunosuppressive responses by inhibiting the activation of CD4+ and CD8+ T cells.^13^ Another study on head and neck squamous cell carcinoma patients found that exoPD-L1 can inhibit the proliferation of CD4+ T cells, induce the apoptosis of CD8+ T cells, and enhance the expression of Treg cells.^14^ These findings suggest that exoPD-L1 is a critical factor in promoting tumor immune evasion, and targeting it could potentially enhance the efficacy of cancer immunotherapy.

ExoPD-L1 is considered the primary mediator of immune escape and tumor progression, as well as a mechanism of treatment resistance. Additionally, circulating exoPD-L1 has been identified as a biomarker for predicting and evaluating the response to immunotherapy.^15^ Recently, Chen et al. found that higher levels of circulating exoPD-L1 prior to anti-PD-1 therapy indicated a poorer prognosis and lower efficacy in melanoma patients. However, after 3-6 cycles of anti-PD-1 therapy, a higher fold change predicted a better prognosis in melanoma patients.^16^ Another study found that the higher the level of circulating exoPD-L1 before surgery in gastric cancer patients, the shorter their overall survival (OS).^17^ Based on the current state of research, it is necessary to conduct a meta-analysis to investigate whether circulating exoPD-L1 is associated with the prognosis of cancer patients. Therefore, we performed this meta-analysis and analyzed the relationship between circulating exoPD-L1 levels and prognosis pre-treatment and post-treatment separately.

## Methods

### Protocol and registration

The recommended reporting criteria for systematic reviews and meta-analyses (PRISMA) statement guidelines were followed when conducting this meta-analysis (Supplemental material 1).^18^ The study protocol was prospectively registered with PROSPERO (CRD42023401030).

### Search strategy

We searched three English-language databases from inception to March 15, 2023, to evaluate studies on the relationship between circulating exoPD-L1 and prognosis in malignant tumors. The three English databases are PubMed, Embase, and Cochrane Library. The following keywords were utilized in the search strategy: “exoPD-L1”, “circulating exosomal PD-L1”, “exosomal PD-L1”, “blood exosomal PD-L1”, “serum exosomal PD-L1”, “plasma exosomal PD-L1”, “cancer”, “tumor”, “malignant tumor”, “malignancies”, “prognosis”, “prognostic”, “outcome”, and “survival”. Only English-language publications were included in this meta-analysis. Reference lists of additional records were manually inspected to identify more candidate articles.

### Inclusion criteria

1. Patients with malignant tumors diagnosed by pathology.
2. Prospective and retrospective studies analyzing the association between circulating exoPD-L1 and survival outcomes in cancer patients.
3. Literature exploring the prognostic value of circulating exoPD-L1 in cancer patients, including OS, recurrence-free survival (RFS), and progression-free survival (PFS). ^19^
4. Studies that directly included hazard ratio (HR) and their 95% confidence interval (CI), or KaplanLJMeier curves.^20^
5. When the results of univariate and multivariate analyses are presented simultaneously in an article, priority should be given to extracting the multivariate data.

### Exclusion criteria

1. Reviews, letters, case reports, or meta-analyses.
2. Studies with incomplete information or insufficient data.
3. Research on animal experiments.

### Quality assessment

The quality of the included studies was assessed using the NewcastleLJOttawa Scale (NOS) in three dimensions: selection, comparability, and results. The NOS quality score ranged from 0 to 9, and studies with a score of 7 or higher were considered high quality, whereas studies with a score lower than 7 were classified as low quality.^21^ Two authors, Qian Cui and Dong Wang, independently assessed the included articles, and the disagreements were resolved by discussion.

### Data extraction

The following items were extracted from each eligible study: the first author’s name, publication year, treatment methods, sample size, cutoff value, time point, and detection methods. Two authors, Wentao Li and Qian Cui, independently extracted data from univariate and multivariate analyses for PFS, RFS, and OS, including the HR and 95% CI. Any controversial issues were resolved through discussion.

### Statistical analysis

All statistical analyses were conducted using STATA 15.1 Software (Stata Corporation, College Station, Texas, USA). The pooled results were calculated using the HR and 95% CI from the included study. Heterogeneity was assessed using Cochran’s q-test and combined *I*^2^ values. P < 0.10 and *I*^2^ ≥ 50% indicate significant heterogeneity in the literature. Subgroup and sensitivity analyses were conducted to explore and interpret the heterogeneity among various types of research (subgroup analyses were performed only for the indicators that included more than four studies). Additionally, the publication bias of the included studies was assessed using Begg’s and Egger’s tests. ^22–23^ A significance level of P < 0.05 was used to determine statistical significance. ^24^

## Results

### Baseline Characteristics of Eligible Studies

A total of 405 studies were obtained following a search through PubMed, Embase, and Cochrane Library. Of these, 377 studies were excluded due to duplications, animal experiments, and reviews. The full-text studies of the remaining 28 articles were then thoroughly evaluated, and 17 studies were removed, leaving 11 studies that were finally included in this meta-analysis. ^16–17, 25–34^ The selection flowchart is displayed in Figure 1.

**Figure 1.**
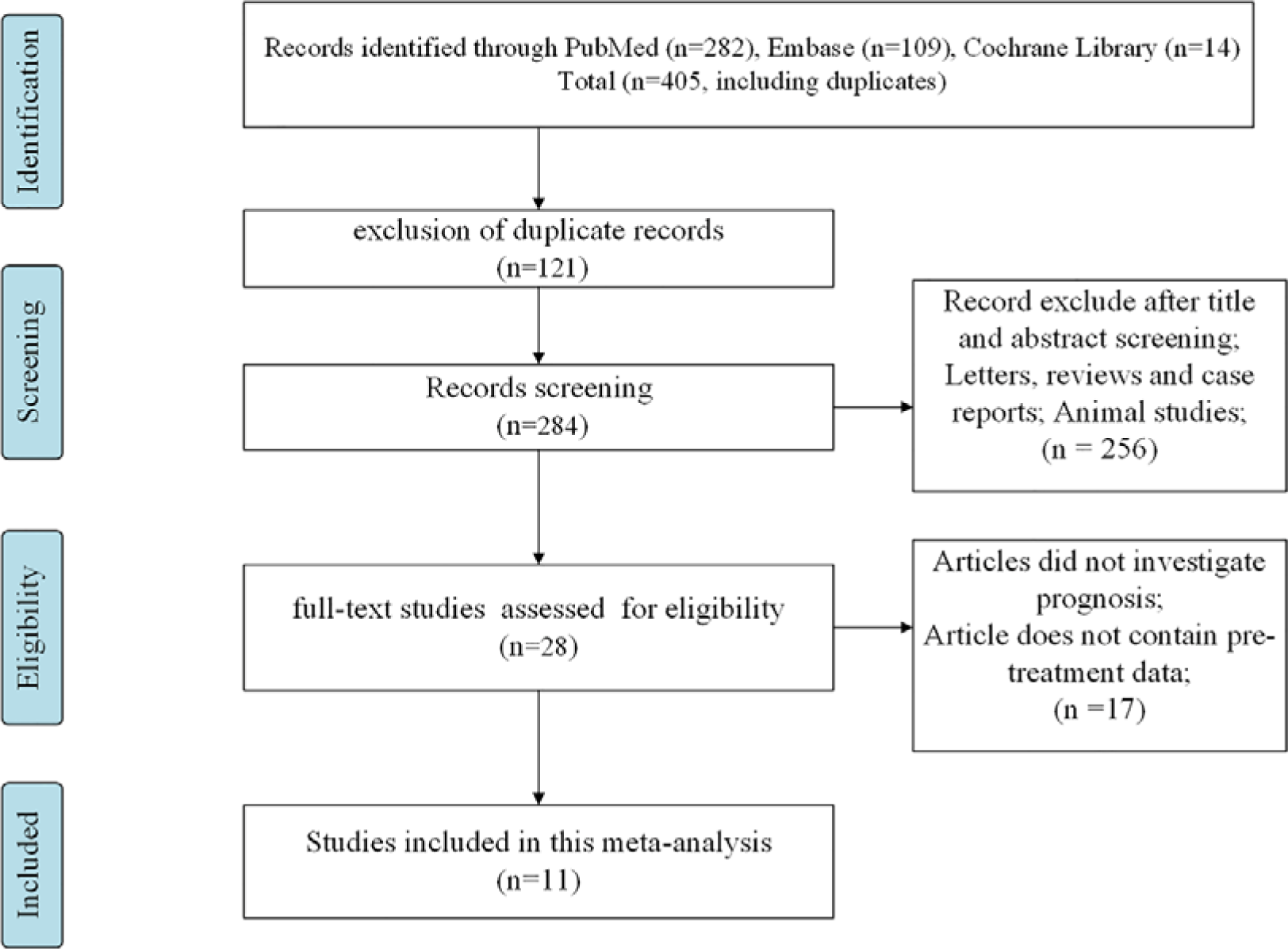
PRISMA flowchart for screening the literature.

As shown in Table 1, all enrolled studies were published between 2018 and 2023. A total of 964 patients with various types of cancer were evaluated, including 360 with lung cancer, 23 with melanoma, 69 with gastric cancer, 55 with pancreatic ductal adenocarcinoma, 107 with extranodal NK/T-cell lymphoma, 62 with castration-resistant prostate cancer, 177 with colorectal liver metastasis, 67 with osteosarcoma, and 44 with various solid tumors. Among the included articles, nine studies investigated the level of pre-treatment exoPD-L1, while three studies included the fold change of exoPD-L1. Four studies used ICIs, while seven articles performed other therapies (chemotherapy, surgery, initial treatment, and new hormonal agents). The quality assessment scores for ten studies were ≥ 7, while 1 study received a score of < 7 (Supplemental Table 1).

**Table 1.**
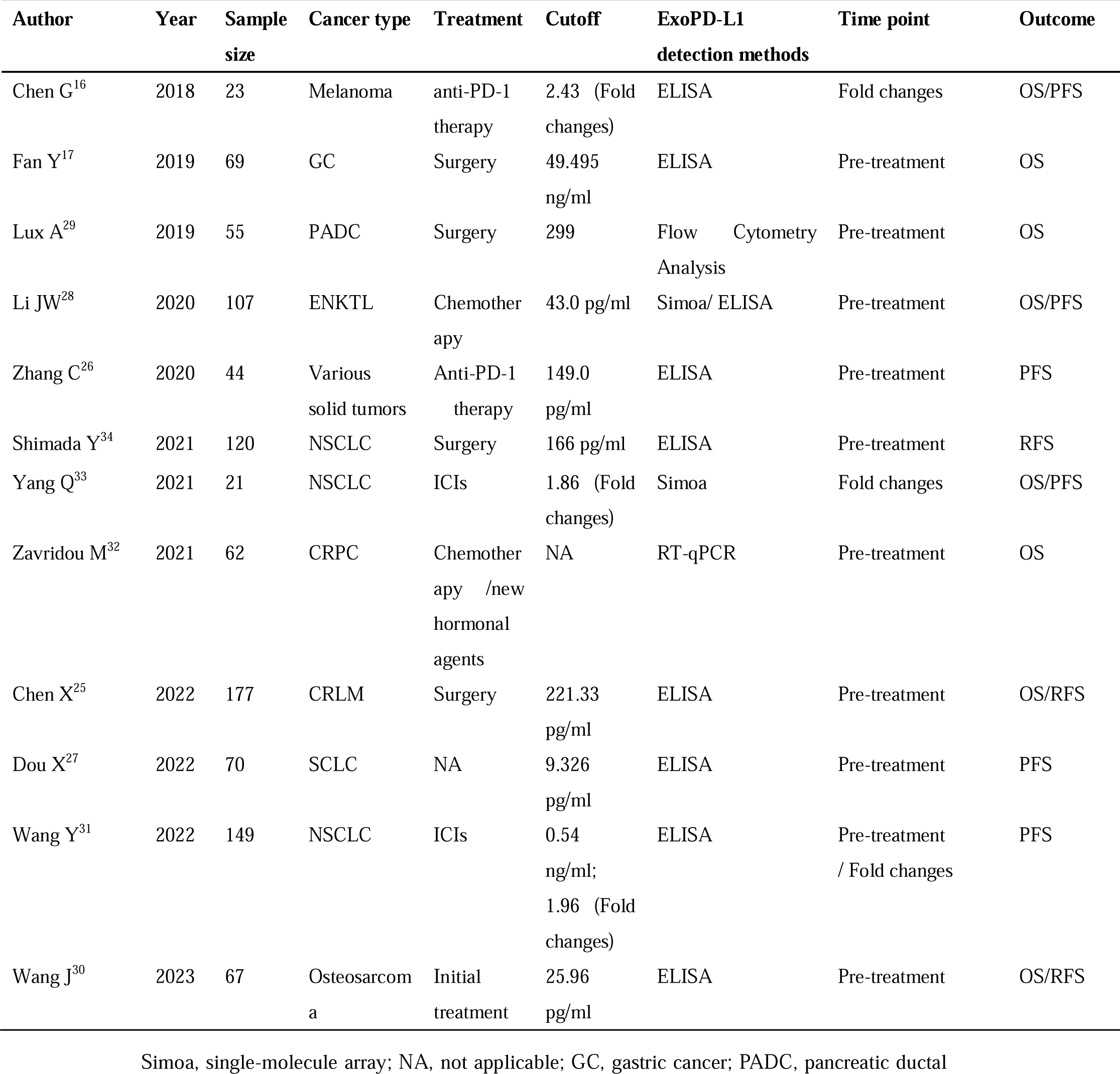
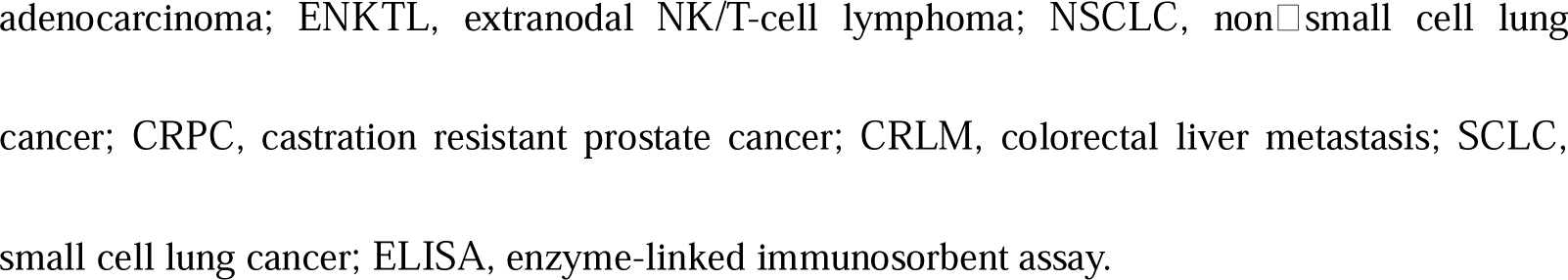
Baseline features of 11 studies.

| Author | Year | Sample size | Cancer type | Treatment | Cutoff | ExoPD-L1 detection methods | Time point | Outcome |
| --- | --- | --- | --- | --- | --- | --- | --- | --- |
| Chen G<sup>16</sup> | 2018 | 23 | Melanoma | anti-PD-1 therapy | 2.43 (Fold changes) | ELISA | Fold changes | OS/PFS |
| Fan Y<sup>17</sup> | 2019 | 69 | GC | Surgery | 49.495 ng/ml | ELISA | Pre-treatment | OS |
| Lux A<sup>29</sup> | 2019 | 55 | PADC | Surgery | 299 | Flow Cytometry Analysis | Pre-treatment | OS |
| Li JW<sup>28</sup> | 2020 | 107 | ENKTL | Chemotherapy | 43.0 pg/ml | Simoa/ ELISA | Pre-treatment | OS/PFS |
| Zhang C<sup>26</sup> | 2020 | 44 | Various solid tumors | Anti-PD-1 therapy | 149.0 pg/ml | ELISA | Pre-treatment | PFS |
| Shimada Y<sup>34</sup> | 2021 | 120 | NSCLC | Surgery | 166 pg/ml | ELISA | Pre-treatment | RFS |
| Yang Q<sup>33</sup> | 2021 | 21 | NSCLC | ICIs | 1.86 (Fold changes) | Simoa | Fold changes | OS/PFS |
| Zavridou M<sup>32</sup> | 2021 | 62 | CRPC | Chemotherapy /new hormonal agents | NA | RT-qPCR | Pre-treatment | OS |
| Chen X<sup>25</sup> | 2022 | 177 | CRLM | Surgery | 221.33 pg/ml | ELISA | Pre-treatment | OS/RFS |
| Dou X<sup>27</sup> | 2022 | 70 | SCLC | NA | 9.326 pg/ml | ELISA | Pre-treatment | PFS |
| Wang Y<sup>31</sup> | 2022 | 149 | NSCLC | ICIs | 0.54 ng/ml; 1.96 (Fold changes) | ELISA | Pre-treatment / Fold changes | PFS |
| Wang J<sup>30</sup> | 2023 | 67 | Osteosarcoma | Initial treatment | 25.96 pg/ml | ELISA | Pre-treatment | OS/RFS |

Simoa, single-molecule array; NA, not applicable; GC, gastric cancer; PADC, pancreatic ductal
adenocarcinoma; ENKTL, extranodal NK/T-cell lymphoma; NSCLC, non-small cell lung cancer; CRPC, castration resistant prostate cancer; CRLM, colorectal liver metastasis; SCLC, small cell lung cancer; ELISA, enzyme-linked immunosorbent assay.

### Effect of pre-treatment exoPD-L1 on prognosis

Figure 2A shows that six studies, comprising 537 cases, investigated the relevance between pre-treatment exoPD-L1 and OS. The pooled data indicate that high concentrations of pre-treatment exoPD-L1 in blood were associated with worse OS than low exoPD-L1 levels (HR = 2.10, 95% CI 1.51–2.91, P < 0.001). There was no significant heterogeneity (*I*^2^ = 0%, P = 0.921), and the pooled results were consistent with the outcomes of the five included studies. ^17, 25, 28–30^.

**Figure 2.**
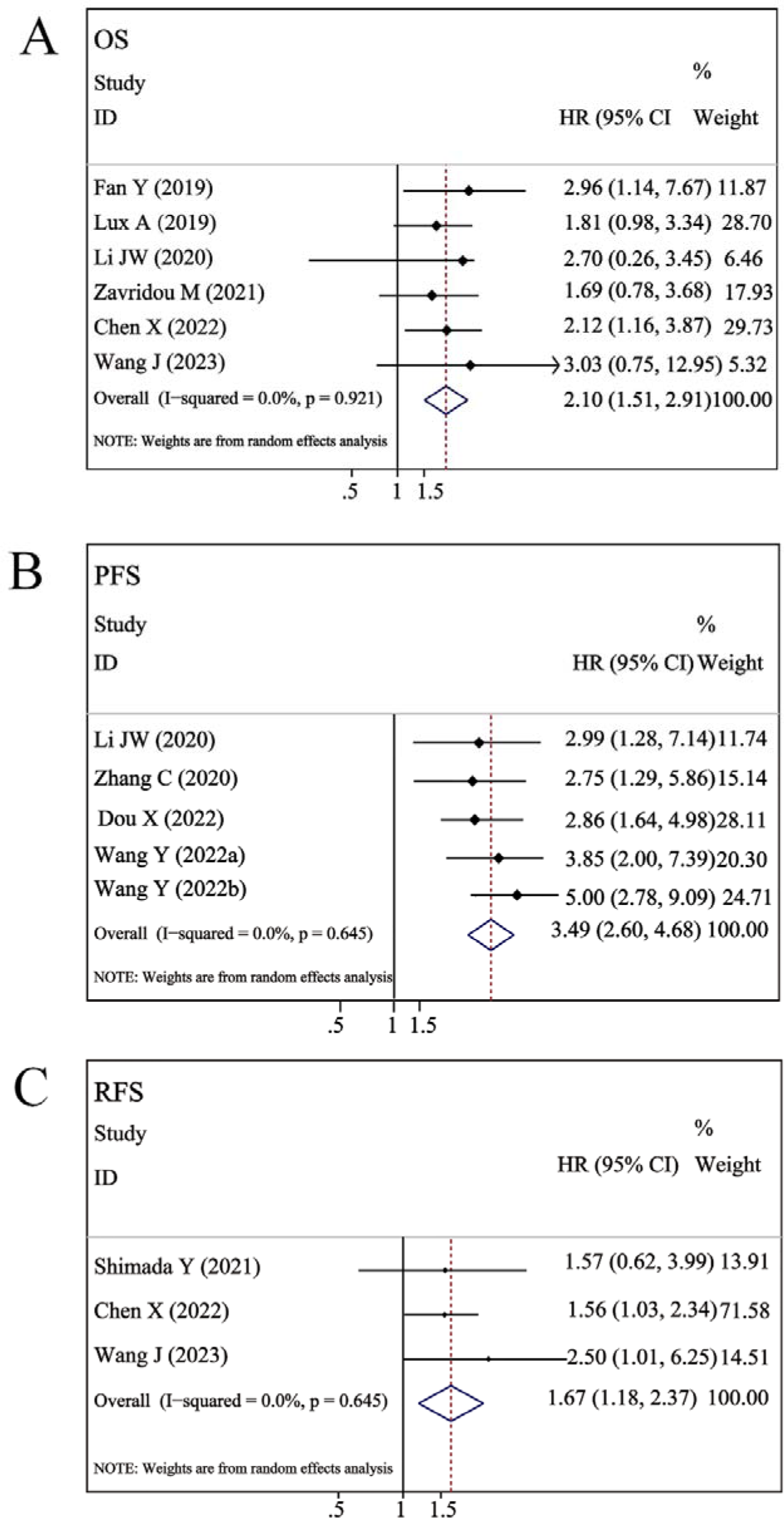
Forest plots of the associations between pre-treatment exoPD-L1 and prognosis. (A) pre-treatment exoPD-L1 versus OS. (B) pre-treatment exoPD-L1 versus PFS. (C) pre-treatment exoPD-L1 versus RFS.

As shown in Figure 2B, four studies, comprising 370 patients, explored the relationship between pre-treatment exoPD-L1 and PFS. Similar to OS, the pooled results indicate that elevated exoPD-L1 levels were correlated with inferior PFS (HR = 3.49, 95% CI 2.60–4.68, P < 0.001). Significant heterogeneity was not found among studies (*I*^2^ = 0%, P = 0.645), and the combined results were consistent with those from the four included studies. ^26–28, 31^

As shown in Figure 2C, three studies, comprising 288 patients, explored the relationship between pre-treatment exoPD-L1 and RFS. The pooled results indicate that elevated exoPD-L1 levels were correlated with inferior RFS (HR = 1.67, 95% CI 1.18–2.37, P < 0.01). Significant heterogeneity was not found among studies (*I*^2^ = 0%, P = 0.645), and the combined results were consistent with those from the three included studies. ^25, 30,34^

In Supplemental Figure 1, removing the studies one by one in sensitivity analyses had no substantial effect on the pooled results, which indicates that the sensitivity analysis was robust.

We conducted subgroup analyses based on tumor types, median cutoff values, sample sizes, and treatment methods to differentiate potential heterogeneity among the included studies. As shown in Table 2, the pooled results for all subgroups were not significantly altered due to low levels of heterogeneity. During the subgroup analysis stratified by tumor type and OS, high levels of exoPD-L1 were significantly associated with poor OS (HR = 2.06, 95% CI 1.47–2.89, P < 0.001) in the subgroup of solid tumors. In a subgroup of hematological malignancies, no statistically significant differences were observed between elevated levels of exoPD-L1 and OS (HR = 2.70, 95% CI 0.74–9.84, P = 0.132; Table 2). In terms of PFS, high levels of exoPD-L1 were significantly associated with poor PFS, whether in solid tumors or hematologic malignancies (solid tumors: HR = 3.56, 95% CI 2.60–4.87, P < 0.001; hematological tumors: HR = 2.99, 95% CI 1.27–7.06, P < 0.05; P _interaction_ = 0.708; Table 2). The stratified analysis of the cutoff value and PFS showed that a higher exoPD-L1 appeared to be linked to a higher HR, but it did not achieve statistical significance (cutoff > 231.33: HR = 4.44; cutoff ≤ 231.33: HR = 2.86; P _interaction_ = 0.145; Table 2). However, this trend was not observed in the subgroup analysis of cutoff value and OS (cutoff > 231.33: HR = 2.09; cutoff ≤ 1: HR = 2.30; P _interaction_ = 0.810; Table 2). Regardless of the treatment method, high circulating exoPD-L1 was significantly associated with shorter PFS (ICIs: HR = 3.94, 95% CI 2.69–5.76, P< 0.001; other therapies: HR =2.90, 95% CI 1.82–4.62, P< 0.001), both in the ICI group and in the other therapy group. The results of the stratified analysis also showed no statistically significant difference between the ICI group and the other therapy group (P _interaction_ = 0.318). However, in the stratified analysis of treatment methods and OS, none of the six studies included treatment with ICIs and only analyzed cancer patients receiving other therapies. The results showed that high levels of exoPD-L1 in the group receiving other therapies predicted poorer OS (HR =2.10, 95% CI 1.51–2.91, P< 0.001) for cancer patients.

**Table 2.** Subgroup analysis of exoPD-L1and OS/PFS.

| Subgroup | N | HR (95% CI) | Interaction | Heterogeneity |  |
| --- | --- | --- | --- | --- | --- |
| Pre-treatment |  |  | P-value | I<sup>2</sup>%, P-value | P<sub>interaction</sub> |
| **OS** |  |  |  |  |  |
| Total | 6 | 2.10 (1.51-2.91) | < 0.001 | 0%, 0.921 |  |
| Caner types |  |  |  |  | 0.692 |
| Solid tumors | 5 | 2.06 (1.47-2.89) | < 0.001 | 0%, 0.867 |  |
| Hematological tumors | 1 | 2.70 (0.74-9.84) | 0.132 | - |  |
| **Sample size** |  |  |  |  | 0.807 |
| > 100 | 2 | 2.21 (1.28-3.82) | < 0.01 | 0%, 0.740 |  |
| ≤ 100 | 4 | 2.03 (1.35-3.06) | < 0.001 | 0%, 0.740 |  |
| **Cut-off value** |  |  |  |  | 0.810 |
| > 221.33 pg/ml | 2 | 2.09 (1.25-3.50) | < 0.01 | 0%, 0.395 |  |
| ≤ 221.33 pg/ml | 3 | 2.30 (1.38-3.84) | < 0.01 | 0%, 0.872 |  |
| NA | 1 | 1.69 (0.78-3.68) | 0.185 |  |  |
| **Treatment methods** |  |  |  |  | - |
| Other therapies | 6 | 2.10 (1.51-2.91) | < 0.001 | 0%, 0.921 |  |
| **Pre-treatment PFS** |  |  |  |  |  |
| Total | 5 | 3.49 (2.60-4.68) | < 0.001 | 0%, 0.645 |  |
| **Caner types** |  |  |  |  | 0.708 |
| Solid tumors | 4 | 3.56 (2.60-4.87) | < 0.001 | 0%, 0.501 |  |
| Hematological tumors | 1 | 2.99 (1.27-7.06) | < 0.05 | - |  |
| **Sample size** |  |  |  |  |  |
| > 100 | 1 | 2.99 (1.27-7.06) | < 0.05 | - | 0.708 |
| ≤ 100 | 4 | 3.56 (2.60-4.87) | < 0.001 | 0%, 0.501 |  |
| **Cut-off value** |  |  |  |  | 0.145 |
| > 221.33 pg/ml | 2 | 4.44 (2.87-6.89) | < 0.001 | 0%, 0.561 |  |
| ≤ 221.33 pg/ml | 3 | 2.86 (1.92-4.25) | < 0.001 | 0%, 0.990 |  |
| **Treatment methods** |  |  |  |  | 0.318 |
| ICIs | 3 | 3.94 (2.69-5.76) | < 0.001 | 0%, 0.474 |  |
| Other therapies | 2 | 2.90 (1.82-4.62) | < 0.001 | 0%, 0.932 |  |
| **Fold change** |  |  |  |  |  |
| **PFS** |  |  |  |  |  |
| Total | 4 | 0.35 (0.23-0.52) | < 0.001 | 0%, 0.482 |  |
| **Caner types** |  |  |  |  |  |
| Solid tumors | 4 | 0.35 (0.23-0.52) | < 0.001 | 0%, 0.482 |  |
| **Sample size** |  |  |  |  | 0.143 |
| > 100 | 2 | 0.40 (0.25-0.63) | < 0.001 | 0%, 0.636 |  |
| ≤ 100 | 2 | 0.18 (0.07-0.48) | < 0.01 | 0%, 0.654 |  |
| **Cut-off value** |  |  |  |  | 0.229 |
| > 1.96 | 1 | 0.14 (0.03-0.63) | < 0.05 | - |  |
| ≤ 1.96 | 3 | 0.37 (0.24-0.58) | < 0.001 | 0%, 0.617 |  |
| **Treatment methods** |  |  |  |  | - |
| ICIs | 4 | 0.35 (0.23-0.52) | < 0.001 | 0%, 0.482 |  |

### Effect of post-treatment fold change in exoPD-L1 on prognosis

In Figure 3A, two studies with 44 cases included the association between post-treatment fold change of exoPD-L1 and OS. The results suggested that high fold changes of circulating exoPD-L1 at post-treatment were significantly correlated with superior OS compared to low fold changes in circulating exoPD-L1 (HR = 0.19, 95% CI 0.10–0.38, P < 0.001), and there was no significant heterogeneity observed among studies (*I*^2^ = 0%, P = 0.625); the pooled results were consistent with the outcomes of the two studies included.^16, 33^

**Figure 3.**
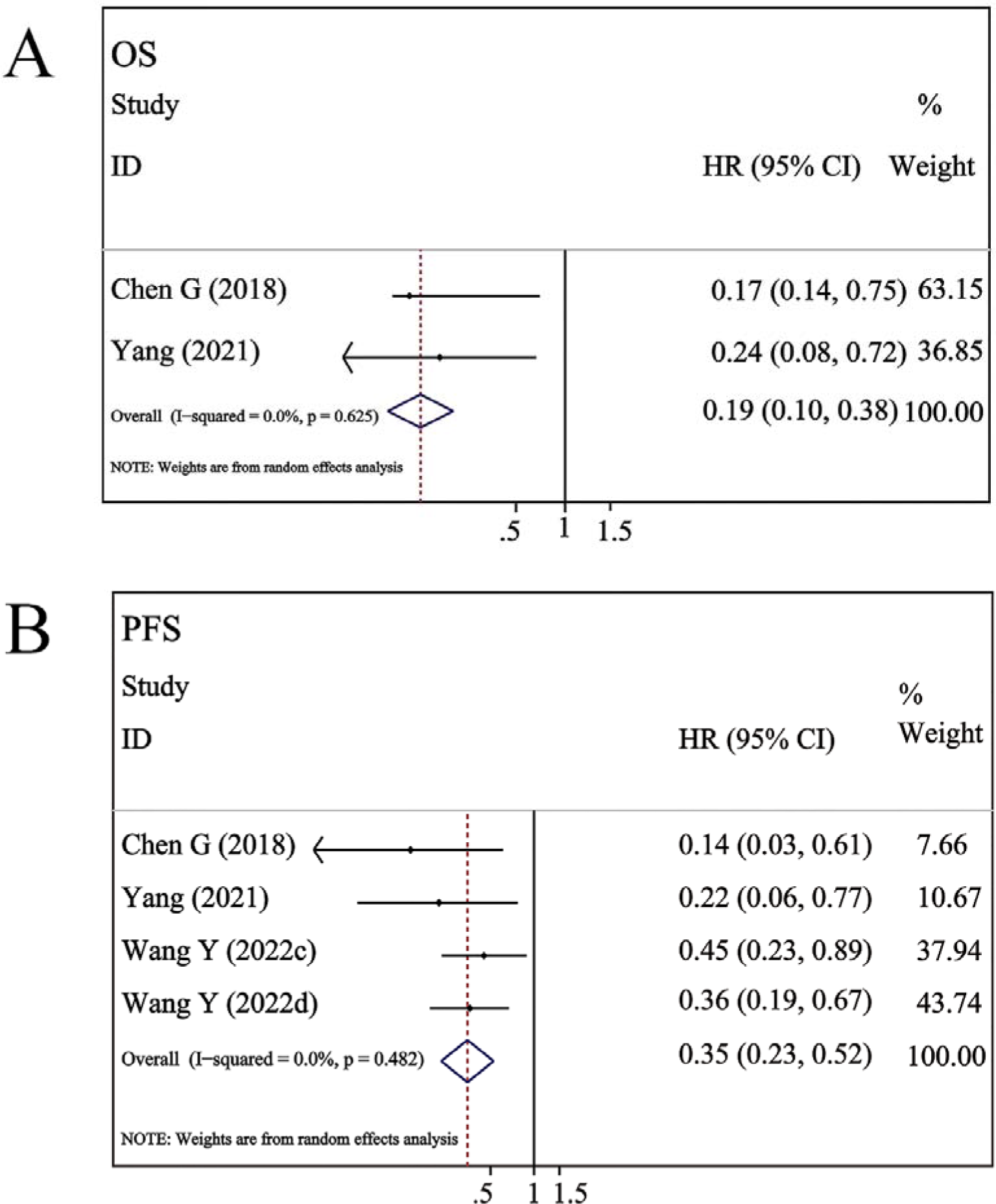
Forest plots of the associations between fold change of exoPD-L1 at post-treatment and prognosis. (A) post-treatment fold change of exoPD-L1 versus OS. (B) post-treatment fold change of exoPD-L1 versus PFS. Wang Y (2022c) received ICI monotherapy. Wang Y (2022d) received ICI combination therapy.

In Figure 3B, three studies comprising 193 patients provided data on the fold change of exoPD-L1 and PFS. Similarly, the pooled analysis of these studies indicated that elevated fold changes of circulating exoPD-L1 after treatment exerted a favorable impact on PFS (HR = 0.35, 95% CI 0.23–0.52, P < 0.001), with no heterogeneity observed (*I*^2^ = 0%, P = 0.482), and the pooled results were consistent with the outcomes of the three studies included.^16, 31, 33^

Since the study investigating the correlation between the fold change of exoPD-L1 and OS included two studies, we could not conduct a subgroup analysis. However, we did conduct a subgroup analysis on the relationship between the fold change of exoPD-L1 and PFS. It is important to note that more studies are necessary to confirm the results presented in this paper. The stratified analysis of the median cutoff value and PFS suggests that a higher fold change of exoPD-L1 may be correlated with a lower HR, although this relationship did not reach statistical significance (cutoff > 1.96: HR = 0.14; cutoff ≤ 1.96: HR = 0.37; P _interaction_ = 0.229; Table 2). Regarding the treatment approach, all three studies used ICIs for treatment. The results showed that a higher fold change of exoPD-L1 at post-treatment among cancer patients who received immune therapy had a longer PFS. Although we did not perform a subgroup analysis of OS and fold change in circulating exoPD-L1, we found that both studies related to OS used ICIs as therapeutic agents (the results are shown in Figure 3A). Through the above analysis, we can also see that current studies on exoPD-L1 after immunotherapy focus on the analysis of its fold change and its correlation with prognosis.

### Publication bias

We assessed publication bias using Begg’s and Egger’s linear regression methods. There was no evidence of publication bias in the included studies (P > 0.05; Supplemental Table 2). The plots for Begg’s and Egger’s tests are shown in Supplemental Figure 2.

## Discussion

Recently, many studies have shown a relationship between exoPD-L1 expression and the prognosis of cancer patients. Some literature indicates that cancer patients with high levels of exoPD-L1 have a poorer prognosis. However, other studies have found that patients with an increasing change in exoPD-L1 can benefit from treatment with immune checkpoint inhibitors. For indicators with divergent results, a meta-analysis can be conducted to guide clinical treatment. However, there is no meta-analysis on the correlation between exoPD-L1 and prognosis. For the first time, we conducted a comprehensive meta-analysis to investigate the prognostic value of circulating exoPD-L1 in various cancer patients. We aim to provide robust evidence on whether circulating exoPD-L1 can serve as an indicator for prognostic evaluation.

In this meta-analysis, we found that high expression of circulating exoPD-L1 in patients before treatment or at baseline was associated with poor PFS, RFS, and OS. However, high fold changes of circulating exoPD-L1 post-treatment were correlated with significantly superior PFS and OS. In the subgroup analysis, we found that high levels of circulating exoPD-L1 before treatment were significantly associated with shorter PFS, regardless of whether the patients received ICIs or other therapies. However, after immunotherapy, high fold changes in circulating exoPD-L1 indicated longer PFS and OS in cancer patients. Our results revealed that exoPD-L1 might serve as an indicator for assessing the prognosis of cancer patients.

Exosomes are extracellular vesicles present in bodily fluids such as blood and urine. talk with the tumor cell microenvironment.^35^ Tumor cells can utilize this mechanism to evade immune responses and promote cancer progression and metastasis.^36^ They can secrete a large amount of PD-L1 within extracellular vesicles. PD-L1 in extracellular vesicles binds to PD-1 through its extracellular domain. Removal of exoPD-L1 can inhibit tumor growth.^37^ Chen et al. found that exoPD-L1 can interact with activated T-cell.^16^ In another study, exoPD-L1 was found to terminate T-cell activation and maintain T-cell exhaustion.^38^ Recent evidence suggests that there may be a negative correlation between the expression levels of circulating exoPD-L1 and CD28 in CD8+ T cells in various advanced tumors.^39^ Notably, several recent studies have found a correlation between the levels of exoPD-L1 isolated from plasma and the status of PD-L1 in tissue.^40–41^ Based on the above analysis, we found that circulating exoPD-L1 is closely associated with cancer progression and immune suppression in cancer patients. Some reviews also mention this view.^12, 42^ Meanwhile, our study found that high levels of exoPD-L1 before treatment were associated with a poor prognosis in cancer patients. These studies further confirm our conclusion.

Previously, we mentioned that exoPD-L1 is considered a central mediator of immune escape and tumor progression.^15^ It is now known that exoPD-L1 has the same affinity as anti-PD-L1 monoclonal antibodies. Circulating exoPD-L1 has been proposed to predict and assess responses to immunotherapy. Reports have indicated that lung cancer patients who are exoPD-L1-positive have better clinical responses to ICIs.^41^ In addition, studies have suggested that patients with elevated exoPD-L1 levels before treatment are in a state of T-cell exhaustion, which may be one of the reasons for poor subsequent immune therapy efficacy. Responders exhibit a significant increase in circulating exoPD-L1 levels 3-6 weeks after treatment. The increase in extracellular exoPD-L1 levels at 3-6 weeks may reflect the successful induction of anti-tumor immunity by immunotherapy. ^16^ Interestingly, several studies have also confirmed an increase in tissue PD-L1 expression in the early stages of treatment in patients who respond to ICIs. ^43–44^

Through the research mentioned above, we found that during the early stages of immunotherapy, PD-L1 expression in tissues and exoPD-L1 in circulation may be upregulated. This could be because immunotherapy blocks the interaction between PD-1 and PD-L1, leading to the release of most of the PD-L1 in exosomes. Therefore, increased PD-L1 expression after receiving immunotherapy may have a positive immune regulatory effect.

### Limitation

There are several limitations to the current meta-analysis that should be acknowledged. First, the number of included studies and patients was small, and more studies with larger samples are needed in the future. Secondly, the included studies used different cutoff values, making direct comparison difficult.

### Conclusion

Overall, our findings indicate that high levels of circulating exoPD-L1 prior to treatment (ICIs and other therapies) are associated with a poor prognosis. However, a high fold change in circulating exoPD-L1 after receiving immunotherapy was correlated with a superior prognosis. Detecting circulating exoPD-L1 through liquid biopsy has the advantages of portability, minimal invasiveness, and repeatability, and it may have important clinical significance for assessing the prognosis of cancer patients.

## Data Availability Statement

All relevant data are within the article and its Supporting Information files.

## Author contributions

Q Cui designed and drafted the manuscript; WT Li and D Wang searched articles; SC Wang checked grammar; JC Yu designed the manuscript.

## Ethical approval and informed consent

Not applicable.

## Consent to publish

Not applicable.

## Acknowledgments

Not applicable.

## Supplemental material

Supplemental material for this article is available online

## References

1. Sun C, Mezzadra R, Schumacher TN. Regulation and function of the PD-L1 checkpoint. Immunity 2018; 48: 434–452.

2. Wu Y, Chen W, Xu ZP, et al. PD-L1 Distribution and Perspective for Cancer Immunotherapy-Blockade, Knockdown, or Inhibition. Front Immunol 2019; 10: 2022.

3. Agata Y, Kawasaki A, Nishimura H, et al. Expression of the PD-1 antigen on the surface of stimulated mouse T and B lymphocytes. Int Immunol 1996; 8: 765–772.

4. Keir ME, Liang SC, Guleria I, et al. Tissue expression of PD-L1 mediates peripheral T cell tolerance. J Exp Med 2006; 203: 883–895.

5. Hui E, Cheung J, Zhu J, et al. T cell costimulatory receptor CD28 is a primary target for PD-1-mediated inhibition. Science 2017; 355: 1428–1433.

6. Page DB, Postow MA, Callahan MK, et al. Immune modulation in cancer with antibodies. Annu Rev Med 2014; 65:185–202.

7. Abril-Rodriguez G, Ribas A. SnapShot: immune checkpoint inhibitors. Cancer Cell 2017; 31: 848–848.

8. Mok TSK, Wu YL, Kudaba I, et al. Pembrolizumab versus chemotherapy for previously untreated, PD-L1-expressing, locally advanced or metastatic non-small-cell lung cancer (KEYNOTE-042): a randomised, open-label, controlled, phase 3 trial. Lancet 2019; 393: 1819-1830.

9. Chen Y, Wen S, Xia J, et al. Association of Dynamic Changes in Peripheral Blood Indexes with Response to PD-1 Inhibitor-Based Combination Therapy and Survival Among Patients with Advanced Non-Small Cell Lung Cancer. Front Immunol 2021; 12: 672271.

10. Rittmeyer A, Barlesi F, Waterkamp D, et al. Atezolizumab versus docetaxel in patients with previously treated non-small-cell lung cancer (OAK): a phase 3, open-label, multicentre randomized controlled trial. Lancet 2017; 389: 255–265.

11. Rashdan S, Minna JD, Gerber DE. Lung cancer immunotherapy biomarker s: refine not reject. Lancet Respir Med 2018; 6: 403.

12. Yin Z, Yu M, Ma T, et al. Mechanisms underlying low-clinical responses to PD-1/PD-L1 blocking antibodies in immunotherapy of cancer: a key role of exosomal PD-L1. J Immunother Cancer 2021; 9: e001698.

13. Ricklefs FL, Alayo Q, Krenzlin H, et al. Immune evasion mediated by PD-L1 on glioblastoma-derived extracellular vesicles. Sci Adv 2018; 4: eaar2766.

14. Theodoraki MN, Yerneni SS, Hoffmann TK, et al. Clinical Significance of PD-L1+ Exosomes in Plasma of Head and Neck Cancer Patients. Clin Cancer Res 2018; 24: 896–905.

15. Daassi D, Mahoney KM, Freeman GJ. The importance of exosomal PDL1 in tumour immune evasion. Nat Rev Immunol 2020; 20: 209–215.

16. Chen G, Huang AC, Zhang W, et al. Exosomal PD-L1 contributes to immunosuppression and is associated with anti-PD-1 response. Nature 2018; 560: 382–386.

17. Fan Y, Che X, Qu J, et al. Exosomal PD-L1 retains immunosuppressive activity and is associated with gastric cancer prognosis. Ann Surg Oncol 2019; 26: 3745–3755.

18. Wang T, Denman D, Bacot SM, Feldman GM. Challenges and the Evolving Landscape of Assessing Blood-Based PD-L1 Expression as a Biomarker for Anti-PD-(L)1 Immunotherapy. Biomedicines 2022; 10: 1181.

19. Mariani P, Russo D, Maisto M, et al. Pretreatment neutrophil-to-lymphocyte ratio is an independent prognostic factor in head and neck squamous cell carcinoma: meta-analysis and trial sequential analysis. J Oral Pathol Med 2022; 51: 39–51.

20. Tierney JF, Stewart LA, Ghersi D, et al. Practical methods for incorporating summary time-to-event data into meta-analysis. Trials 2007; 8: 16.

21. Jiang T, Bai Y, Zhou F, et al. Clinical value of neutrophil-to-lymphocyte ratio in patients with non-small-cell lung cancer treated with PD-1/PD-L1 inhibitors. Lung Cancer 2019; 130: 76–83.

22. Begg CB, Mazumdar M. Operating characteristics of a rank correlation test for publication bias. Biometrics 1994; 50: 1088–1101.

23. Egger M, Davey Smith G, Schneider M, et al. Bias in meta-analysis detected by a simple, graphical test. BMJ 1997; 316: 629–634.

24. Li ML, Luo HY, Quan ZW, et al. Prognostic and clinicopathologic significance of PLIN2 in cancers: A systematic review with meta-analysis. Int J Biol Markers 2023; 38: 3–14.

25. Chen X, Du Z, Huang M, et al. Circulating PD-L1 is associated with T cell infiltration and predicts prognosis in patients with CRLM following hepatic resection. Cancer Immunol Immunother 2022; 71: 661–674.

26. Zhang C, Fan Y, Che X, et al. Anti-PD-1 Therapy Response Predicted by the Combination of Exosomal PD-L1 and CD28. Front Oncol 2020; 10: 760.

27. Dou X, Hua Y, Chen Z, et al. Extracellular vesicles containing PD-L1 contribute to CD8+T-cell immune suppression and predict poor outcomes in small cell lung cancer. Clin Exp Immunol 2022; 207: 307–317.

28. Li JW, Wei P, Guo Y, et al. Clinical significance of circulating exosomal PD-L1 and soluble PD-L1 in extranodal NK/T-cell lymphoma, nasal-type. Am J Cancer Res 2020; 10: 4498–4512.

29. Lux A, Kahlert C, Grützmann R, et al. c-Met and PD-L1 on Circulating Exosomes as Diagnostic and Prognostic Markers for Pancreatic Cancer. Int J Mol Sci 2019; 20: 3305.

30. Wang J, Guo W, Wang X, et al. Circulating Exosomal PD-L1 at Initial Diagnosis Predicts Outcome and Survival of Patients with Osteosarcoma. Clin Cancer Res 2023; 29: 659–666.

31. Wang Y, Niu X, Cheng Y, et al. Exosomal PD-L1 predicts response with immunotherapy in NSCLC patients. Clin Exp Immunol 2022; 208: 316–322.

32. Zavridou M, Strati A, Bournakis E, et al. Prognostic Significance of Gene Expression and DNA Methylation Markers in Circulating Tumor Cells a nd Paired Plasma Derived Exosomes in Metastatic Castration Resistant Prostate Cancer. Cancers 2021; 13: 780.

33. Yang Q, Chen M, Gu J, et al. Novel Biomarkers of Dynamic Blood PD-L1 Expression for Immune Checkpoint Inhibitors in Advanced Non-Small-Cell Lung Cancer Patients. Front Immunol 2021; 12: 665133.

34. Shimada Y, Matsubayashi J, Kudo Y, et al. SerumLJderived exosomal PDLJL1 expression to predict antiLJPDLJ1 response and in patients with nonLJsmall cell lung cancer. Sci Rep 2021; 11: 7830.

35. Boukouris S, Mathivanan S. Exosomes in bodily fluids are a highly stable resource;e of disease biomarkers. Proteomics Clin Appl 2015; 9: 358–367.

36. Chen DS, Mellman I. Elements of cancer immunity and the cancer–immune set point. Nature 2017; 541: 321–330.

37. Poggio M, Hu T, Pai CC, et al. Suppression of Exosomal PD-L1 Induces Systemic Anti-tumor Immunity and Memory. Cell 2019; 177: 414–427.

38. Huang AC, Postow MA, Orlowski RJ, et al. T-cell invigoration to tumour burden ratio associated with anti-PD-1 response. Nature 2017; 545: 60–65.

39. Zhang C, Fan Y, Che X, et al. Anti-PD-1 therapy response predicted by the combination of exosomal PD-L1 and CD28. Front Oncol 2020; 10: 760.

40. Kim DH, Kim H, Choi YJ, et al. Exosomal PD-L1 promotes tumor growth through immune escape in non-small cell lung cancer. Exp Mol Med 2019; 51: 1–13.

41. Zhang Z, Jin W, Xu K, et al. Blood exosome PD-L1 is associated with PD-L1 expression measured by immunohistochemistry, and lymph node metastasis in lung cancer. Tissue Cell 2022; 79: 101941.

42. Xie F, Xu M, Lu J, et al. The role of exosomal PD-L1 in tumor progres sion and immunotherapy. Mol Cancer 2019; 18: 146.

43. Vilain RE, Menzies AM, Wilmott JS, et al. Dynamic changes in PD-L1 expression and immune infiltrates early during treatment predict response to PD-1 blockade in melanoma. Clin Cancer Res 2017; 23: 5024–5033.

44. Chen PL, Roh W, Reuben A, et al. Analysis of immune signatures in longitudinal tumor samples yields insight into biomarkers of response and mechanisms of resistance to immune checkpoint blockade. Cancer Discov 2016; 6: 827–837.

